# Pattern of Standard Modifiable Risk Factors and Outcomes in Very Young Patients with Myocardial Infarction

**DOI:** 10.1101/2024.11.04.24316727

**Authors:** Si-qi Tang, Xin-long Zhao, Quan Li, Yan-bo Liu, Yi-tao Han, Yu-xiong Chen, Jin-yan Lei, Ya-kun Zhao, Zhong-jie Fan, Yan-ping Ruan

## Abstract

**Background:** The incidence of myocardial infarction (MI) is rising among young individuals. This study aimed to identify the profile of standard modifiable risk factors (SMuRFs) and their impact on long-term survival in very young MI patients (≤35 years).

**Methods:** The VYTAL-MI study (Very Young Tracking and Analysis of Long-term Myocardial Infarction Outcomes) is a retrospective cohort analysis of MI patients aged ≤35 years, admitted to Beijing Anzhen Hospital and Peking Union Medical College Hospital between December 1^1920^, 2011, and December 1^1920^, 2021. SMuRFs (hypertension, diabetes, hypercholesterolemia, smoking) were documented. The primary outcome was major adverse cardiovascular and cerebrovascular events (MACCE), including all-cause death, recurrent MI, ischemia-driven revascularization, and ischemic stroke. Cox proportional hazard models were used to assess associations between SMuRFs and outcomes.

**Results:** Among 690 patients (median age 33 years [interquartile range, IQR: 30-34]; 95% male), only 8% had no SMuRFs. Smoking was most common (76%), followed by hypercholesterolemia (43%), hypertension (40%), and diabetes (12%). The most frequent combination was smoking and hypercholesterolemia (16.4%). Over a median follow-up of 6.3 years (IQR: 4.0-8.6), MACCE occurred in 165 patients (23.9%). Patients with ≥3 SMuRFs had significantly higher cumulative hazards (*P*=0.003). Hypertension (hazard ratio, HR: 1.50; 95% confidence interval, CI: 1.09-2.06; *P*=0.014) and diabetes (HR: 1.67; 95% CI: 1.10-2.54; *P*=0.016) were independently associated with increased risk, while hypercholesterolemia (HR: 1.24; 95% CI: 0.91-1.68; *P*=0.177) and smoking (HR: 1.06; 95% CI: 0.72-1.55; *P*=0.776) showed no significant association.

**Conclusions:** Very young MI patients exhibited a high burden of SMuRFs, particularly smoking. Hypertension and diabetes increased long-term event risk. Targeted SMuRF management is essential to improve survival in this population.

**What is known:**

- Targeting standard modifiable cardiovascular risk factors, including hypertension, diabetes, hypercholesterolemia, and smoking, is crucial for improving outcomes in myocardial infarction.
- Limited data exist on the pattern of standard modifiable risk factors in very young myocardial infarction patients aged ≤35 years, and how these factors influence long-term outcomes.

**What the study adds:**

- Very young myocardial infarction patients exhibited a substantial burden of standard modifiable risk factors, with smoking being notably prevalent.
- Increased standard modifiable risk factors exposure was associated with an elevated risk of major adverse cardiovascular and cerebrovascular events
- Hypertension and diabetes were linked to a higher risk of major adverse cardiovascular and cerebrovascular events, while smoking and hypercholesterolemia did not show a significant independent effect.

**Registration:** URL: http://www.chictr.org.cn; Unique identifier: ChiCTR2400085600.

## Introduction

Myocardial infarction (MI) remains a substantial cause of premature cardiovascular death worldwide.^1,2^ Despite advances in preventive therapies, younger individuals have experienced a slower decline in MI incidence and mortality than elder populations.^3^ The Atherosclerosis Risk in Communities (ARIC) Surveillance study reported an increase in the proportion of MI patients aged 35 to 54 years, rising from 27% to 32% between 1995 and 2014.^4^ Similarly, a Chinese cohort observed a 57% increase in MI cases among individuals under 45 years old between 2010 and 2014 in Beijing.^5^ Although non-traditional risk factors, such as autoimmune diseases, sleep apnea, and substance abuse contribute to this trend,^6–8^ the increase is primarily driven by the growing prevalence of modifiable risk factors.^9,10^ Targeting standard modifiable cardiovascular risk factors (SMuRFs), including hypertension, diabetes, hypercholesterolemia, and smoking, has substantially improved the MI outcome.^11^

Premature MI patients have a poor overall prognosis, experiencing frequent recurrences and premature deaths in the following decades.^12^ However, existing data highlighted that premature MI patients have insufficient control of risk factors.^3,13^ Current guidelines for primary and secondary prevention are predominantly age-dependent, often underestimating coronary artery disease (CAD) risk in younger patients.^14,15^ For example, the 10-year Atherosclerotic Cardiovascular Disease risk calculator does not apply to individuals under 40 years. The Duke Databank for Cardiovascular Disease (DDCD) study found that less than half of MI patients under 55 were eligible for statins before their index MI, and only 1/4 met the criteria for very high-risk events.^16^

Previous studies have defined premature MI as the first occurrence of MI before the age of 45 or 50.^12^ Patients under 35 with MI are often underrepresented, comprising less than 20% of studied populations. Very young MI patients may have distinct risk profiles. The DDCD study, which included 239 patients under 35, found that these individuals had a higher body mass index (BMI) but fewer cardiovascular risk factors compared to those aged 35-45.^13^ Similarly, the Partners YOUNG-MI Registry compared 431 patients under 40 with 1,666 patients aged 41-50, revealing lower hypertension rates and higher substance abuse rates in the younger group.^6^ Focused studies on modifiable risk factors and long-term outcomes in very young MI patients remain limited, but are crucial for informing preventive strategies for this population.

The VYTAL-MI study (Very Young Tracking and Analysis of Long-term Myocardial Infarction Outcomes) is a contemporary, double-center Chinese cohort study focused on very young MI patients (≤35 years). This study aimed to identify the prevalence and patterns of standard modifiable cardiovascular risk factors (SMuRFs) and to evaluate their impact on long-term survival.

## Methods

### Study population

The VYTAL-MI study is a retrospective cohort study conducted at Beijing Anzhen Hospital, Capital Medical University, and Peking Union Medical College Hospital, Chinese Academy of Medical Sciences, two tertiary medical centers in Beijing, China. We consecutively included patients diagnosed with MI before the age of 35 years from 1 December 2011 to 1 December 2021, as defined by the Fourth Universal Definition of Myocardial Infarction by the World Health Organization.^17^ Patients were identified based on primary and secondary diagnoses coded according to the 10th revision of the International Classification of Diseases (ICD-10) codes I21–I22. The analysis included patients with angiographically confirmed coronary artery disease (CAD), defined as ≥50% stenosis in at least one epicardial vessel. Patients with coexisting cardiomyopathy, myocarditis, missing coronary angiography, or follow-up duration of less than one year were excluded. All data were collected anonymously. The Institutional Ethics Committee of Beijing Anzhen Hospital, Capital Medical University, and Peking Union Medical College Hospital, Chinese Academy of Medical Sciences approved this study. All subjects provided original consent for research use of their data, and re-consent was waived. The study was registered with the Chinese Clinical Trial Registry (URL: http://www.chictr.org.cn; Unique identifier: ChiCTR2400085600).

### Definition of SMuRFs

SMuRFs included hypertension, diabetes, hypercholesterolemia, and smoking. Hypertension was defined as a previous diagnosis, use of antihypertensive medication, or a new diagnosis during the index admission. Diabetes was determined by a previous diagnosis, use of glucose-lowering medications, or a new diagnosis during the index admission. Hypercholesterolemia was defined by a previous diagnosis, use of low-density lipoprotein cholesterol (LDL-C)-lowering therapy, an LDL-C level greater than 3.5 mmol/L, or a total cholesterol level greater than 5.5 mmol/L during the index admission. Smoking was defined as smoking more than one cigarette per day within 30 days.^18^ All the patients had complete SMuRF data extracted from structured electronic medical records.

### Data collection and follow-up

Data were extracted from the electronic medical record systems of both hospitals. Baseline clinical characteristics included presenting features, cardiovascular risk factors, comorbid conditions, laboratory and echocardiographic findings, angiographic findings, and medications at discharge. A family history of premature CAD was defined as a first-degree relative hospitalized due to MI or angina with coronary revascularization before 55 years of age in men and 65 years in women. Left ventricular ejection fraction (LVEF) was measured using the biplane Simpson method with echocardiography. All angiograms were reviewed for the number and location of vessels with significant stenosis (≥50%). Intravascular ultrasound or optical coherence tomography assisted in determining the possible pathogenesis of MI. Revascularization strategies were recorded, including percutaneous coronary intervention (PCI), coronary artery bypass grafting (CABG), and intravenous thrombolysis. The discharge medication regimen was recorded, encompassing antiplatelet therapy, β-blockers, angiotensin-converting enzyme (ACE) inhibitors or angiotensin receptor blockers (ARBs), and lipid-lowering agents.

Follow-up information was obtained from recommended clinical visit records at 1 month, 3 months, 6 months, 12 months, and annually thereafter. Additional data were collected through phone interviews for all patients between 1 September 2023 and 29 February 2024, with follow-ups censored on 29 February 2024, regardless of event occurrence. Follow-up information included the occurrence of MACCE, self-reported angina, new occurrences of diabetes, smoking status, and current medications. New-onset diabetes was identified based on the self-reported initiation of glycaemic control therapy.

### Endpoint definition

The primary endpoint was a composite of major adverse cardiovascular and cerebrovascular events (MACCE), including all-cause death, recurrent MI, ischemia-driven revascularization, and ischemic stroke. MI was defined according to the Fourth Universal Definition of MI, which includes evidence of myocardial necrosis in a clinical setting consistent with myocardial ischemia.^19^ Ischemia-driven revascularization included any PCI or CABG performed due to documented myocardial ischemia. Ischemic stroke was defined as an acute episode of focal or global neurological dysfunction caused by cerebral infarction, in line with the American Heart Association/American Stroke Association guidelines.

### Statistical Analysis

Differences in baseline characteristics between patients with and without SMuRFs were assessed using the independent t-test for normally distributed continuous variables, presented as mean ± standard deviation (SD), and the Mann-Whitney U test for non-normally distributed continuous variables, presented as median (interquartile range, IQR). Categorical variables were analyzed using either the Chi-Square test or Fisher’s Exact Test, based on expected frequencies in contingency tables. Fisher’s Exact Test was used when any expected frequency was below 5. The distribution and combination of risk factors were visualized using an UpSet plot, generated with the ’UpSetR’ package in R.

Cumulative incidence rates were calculated as the number of new MACCEs per 100 person-years of follow-up. Kaplan-Meier curves were used to estimate cumulative event-free survival for the primary endpoint, stratified by the number of SMuRFs, and compared using the log-rank test. Pairwise log-rank tests were conducted to compare survival among groups. To control for multiple comparisons, the Benjamini-Hochberg adjustment was applied. Associations between exposure variables and MACCE were assessed using multivariate Cox proportional hazards models, with calculated hazard ratios (HRs) and 95% confidence intervals (CIs). Separate models were fitted for each SMuRF (hypertension, diabetes, hypercholesterolemia, and smoking) and combinations of two risk factors (e.g., diabetes and hypertension). The first set of multivariable Cox models adjusted for age, sex, β-blockers, ACE inhibitors/ARBs, and statins at discharge. The second set was adjusted for age, sex, family history of premature CAD, obesity, and the remaining SMuRFs. Results were presented in forest plots. Interaction analyses between SMuRFs were conducted using both unadjusted and adjusted Cox proportional hazards models to assess their combined impact on the risk of MACCE.

The significance level was *P* < 0.05, and all tests were two-sided. All statistical analyses were performed using R statistical software (version 4.3.0).

## Results

### Baseline Characteristics and Outcome

Of the 934 patients identified with MI, three were diagnosed with myocarditis or cardiomyopathy at discharge, 37 were excluded due to missing laboratory records or angiography, 98 were lost to follow-up, and 106 were excluded for lack of significant stenosis (Figure 1). Finally, 690 MI patients with obstructive CAD were analyzed, with a median age of 33 years (IQR: 30-34), and the majority being male (95%). Among the cohort, 62% presented with ST-segment elevation myocardial infarction (STEMI), while only 55 patients (8%) were classified as SMuRF-less. Single-vessel disease was the predominant presentation (54%), with the left anterior descending artery being the most frequently affected vessel (73%). Interventions included PCI in 83% of patients and CABG in 4% (Table 1).

**Figure 1.**
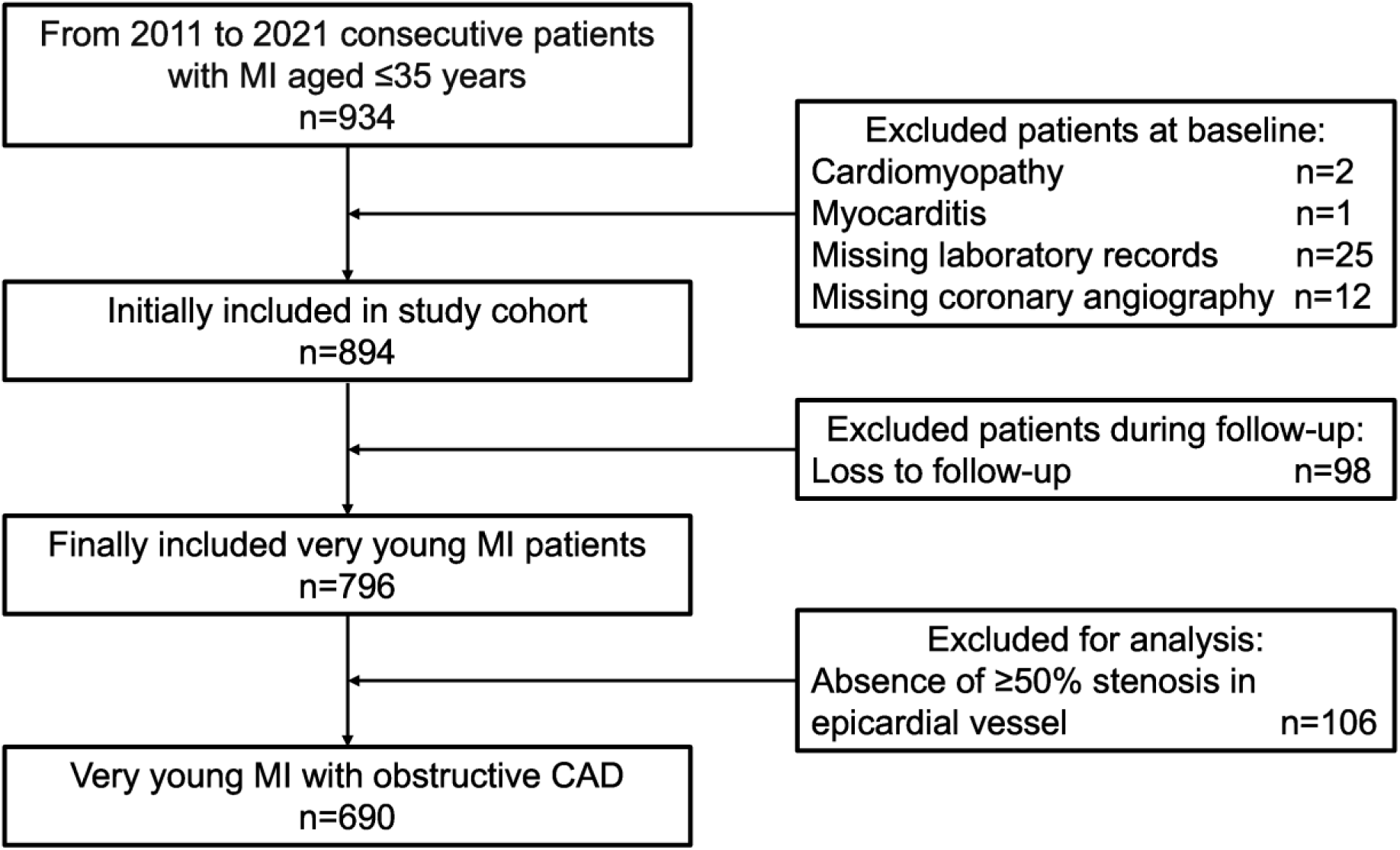
Selection of study population. CAD, coronary artery disease; MI, myocardial infarction

**Table 1.**
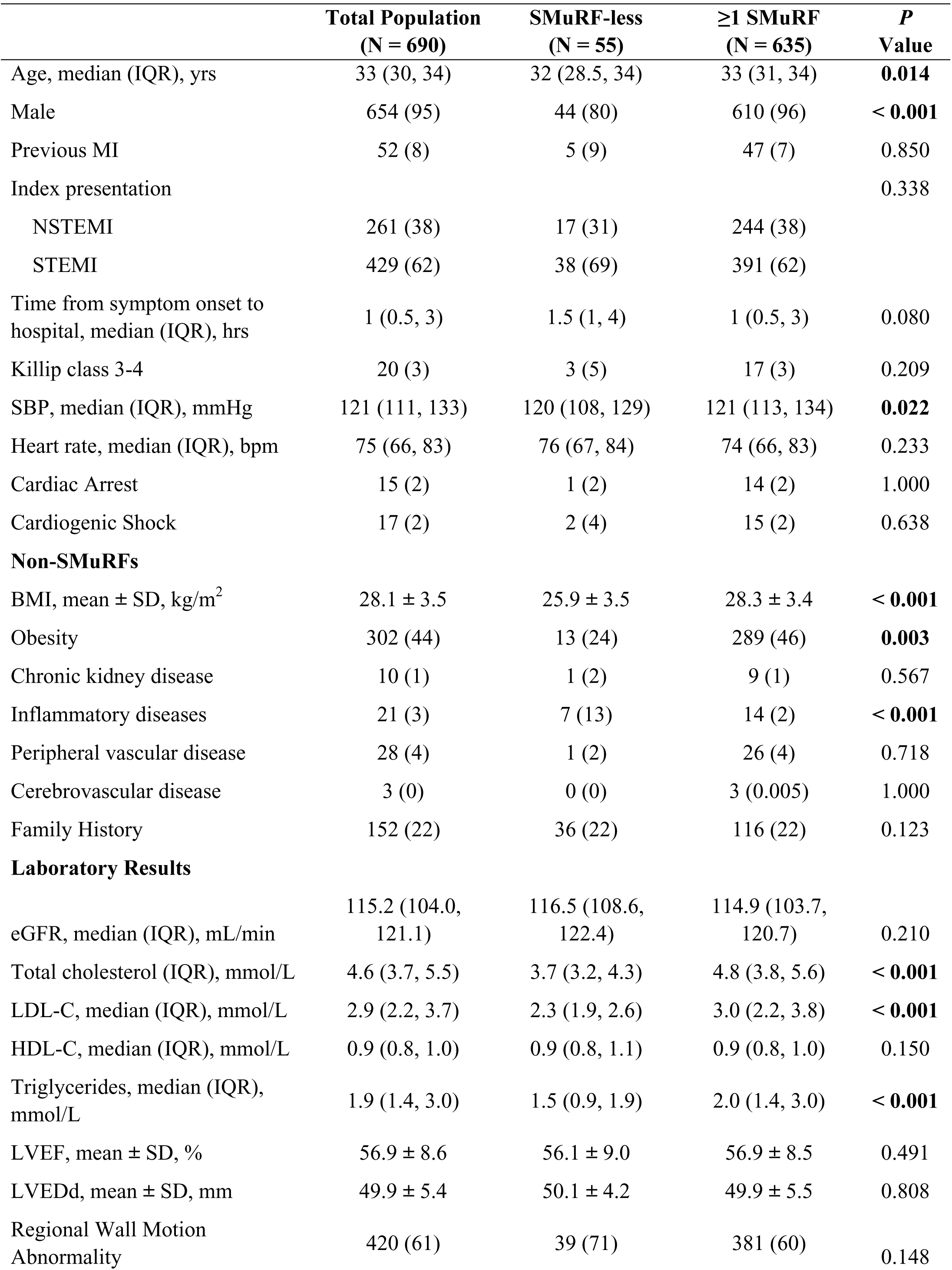
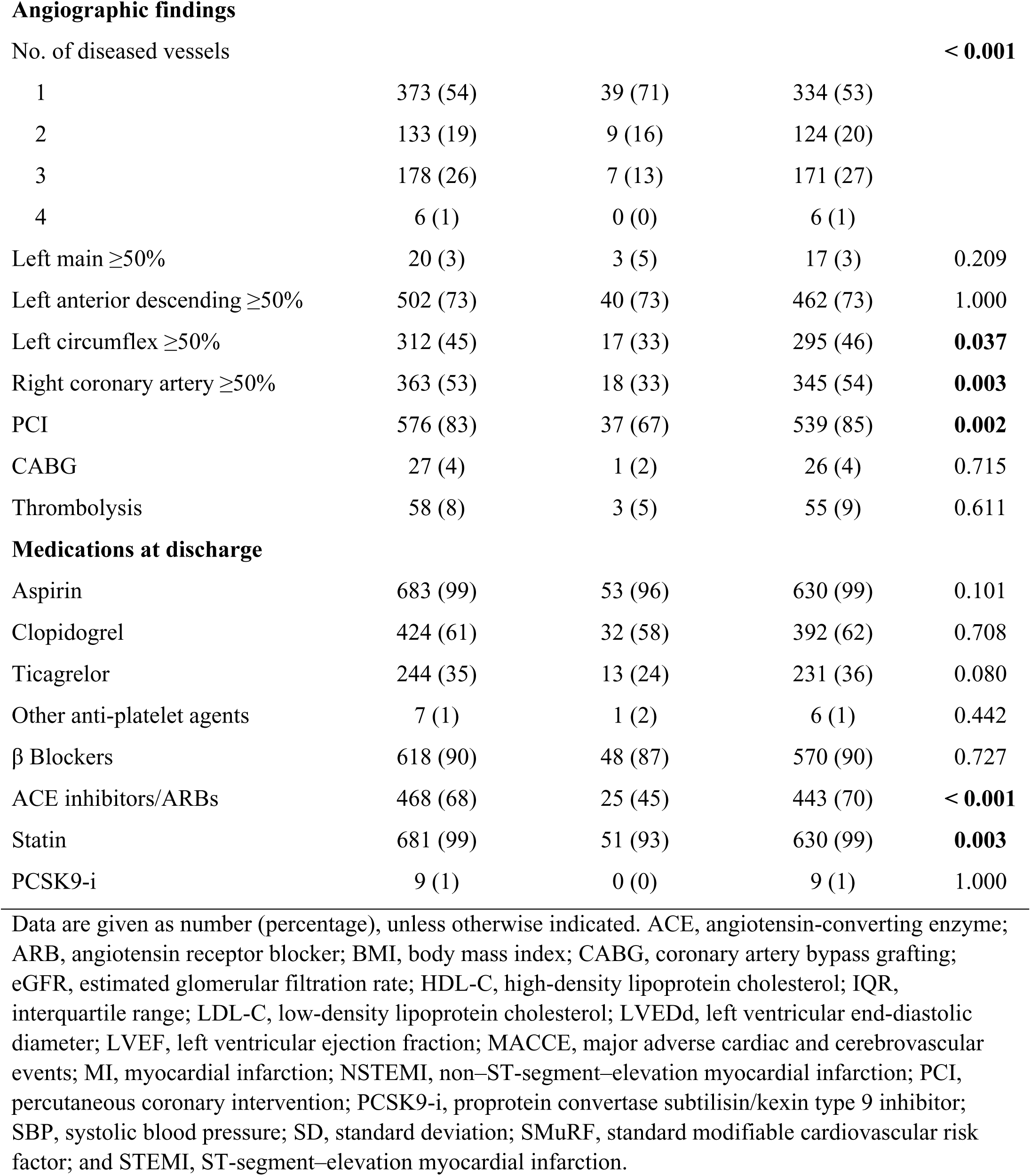
Baseline Patient Characteristics According to With and Without SMuRFs

|  | **Total Population**<br>**(N = 690)** | **SMuRF-less**<br>**(N = 55)** | **≥1 SMuRF**<br>**(N = 635)** | **P**<br>**Value** |
| --- | --- | --- | --- | --- |
| Age, median (IQR), yrs | 33 (30, 34) | 32 (28.5, 34) | 33 (31, 34) | **0.014** |
| Male | 654 (95) | 44 (80) | 610 (96) | **< 0.001** |
| Previous MI | 52 (8) | 5 (9) | 47 (7) | 0.850 |
| Index presentation |  |  |  | 0.338 |
| NSTEMI | 261 (38) | 17 (31) | 244 (38) |  |
| STEMI | 429 (62) | 38 (69) | 391 (62) |  |
| Time from symptom onset to<br>hospital, median (IQR), hrs | 1 (0.5, 3) | 1.5 (1, 4) | 1 (0.5, 3) | 0.080 |
| Killip class 3-4 | 20 (3) | 3 (5) | 17 (3) | 0.209 |
| SBP, median (IQR), mmHg | 121 (111, 133) | 120 (108, 129) | 121 (113, 134) | **0.022** |
| Heart rate, median (IQR), bpm | 75 (66, 83) | 76 (67, 84) | 74 (66, 83) | 0.233 |
| Cardiac Arrest | 15 (2) | 1 (2) | 14 (2) | 1.000 |
| Cardiogenic Shock | 17 (2) | 2 (4) | 15 (2) | 0.638 |
| **Non-SMuRFs** |  |  |  |  |
| BMI, mean ± SD, kg/m<sup>2</sup> | 28.1 ± 3.5 | 25.9 ± 3.5 | 28.3 ± 3.4 | **< 0.001** |
| Obesity | 302 (44) | 13 (24) | 289 (46) | **0.003** |
| Chronic kidney disease | 10 (1) | 1 (2) | 9 (1) | 0.567 |
| Inflammatory diseases | 21 (3) | 7 (13) | 14 (2) | **< 0.001** |
| Peripheral vascular disease | 28 (4) | 1 (2) | 26 (4) | 0.718 |
| Cerebrovascular disease | 3 (0) | 0 (0) | 3 (0.005) | 1.000 |
| Family History | 152 (22) | 36 (22) | 116 (22) | 0.123 |
| **Laboratory Results** |  |  |  |  |
| eGFR, median (IQR), mL/min | 115.2 (104.0,<br>121.1) | 116.5 (108.6,<br>122.4) | 114.9 (103.7,<br>120.7) | 0.210 |
| Total cholesterol (IQR), mmol/L | 4.6 (3.7, 5.5) | 3.7 (3.2, 4.3) | 4.8 (3.8, 5.6) | **< 0.001** |
| LDL-C, median (IQR), mmol/L | 2.9 (2.2, 3.7) | 2.3 (1.9, 2.6) | 3.0 (2.2, 3.8) | **< 0.001** |
| HDL-C, median (IQR), mmol/L | 0.9 (0.8, 1.0) | 0.9 (0.8, 1.1) | 0.9 (0.8, 1.0) | 0.150 |
| Triglycerides, median (IQR),<br>mmol/L | 1.9 (1.4, 3.0) | 1.5 (0.9, 1.9) | 2.0 (1.4, 3.0) | **< 0.001** |
| LVEF, mean ± SD, % | 56.9 ± 8.6 | 56.1 ± 9.0 | 56.9 ± 8.5 | 0.491 |
| LVEDd, mean ± SD, mm | 49.9 ± 5.4 | 50.1 ± 4.2 | 49.9 ± 5.5 | 0.808 |
| Regional Wall Motion<br>Abnormality | 420 (61) | 39 (71) | 381 (60) | 0.148 |
| Angiographic findings |  |  |  |  |
| No. of diseased vessels |  |  |  | **< 0.001** |
| 1 | 373 (54) | 39 (71) | 334 (53) |  |
| 2 | 133 (19) | 9 (16) | 124 (20) |  |
| 3 | 178 (26) | 7 (13) | 171 (27) |  |
| 4 | 6 (1) | 0 (0) | 6 (1) |  |
| Left main $\geq 50\%$ | 20 (3) | 3 (5) | 17 (3) | 0.209 |
| Left anterior descending $\geq 50\%$ | 502 (73) | 40 (73) | 462 (73) | 1.000 |
| Left circumflex $\geq 50\%$ | 312 (45) | 17 (33) | 295 (46) | **0.037** |
| Right coronary artery $\geq 50\%$ | 363 (53) | 18 (33) | 345 (54) | **0.003** |
| PCI | 576 (83) | 37 (67) | 539 (85) | **0.002** |
| CABG | 27 (4) | 1 (2) | 26 (4) | 0.715 |
| Thrombolysis | 58 (8) | 3 (5) | 55 (9) | 0.611 |
| **Medications at discharge** |  |  |  |  |
| Aspirin | 683 (99) | 53 (96) | 630 (99) | 0.101 |
| Clopidogrel | 424 (61) | 32 (58) | 392 (62) | 0.708 |
| Ticagrelor | 244 (35) | 13 (24) | 231 (36) | 0.080 |
| Other anti-platelet agents | 7 (1) | 1 (2) | 6 (1) | 0.442 |
| $\beta$ Blockers | 618 (90) | 48 (87) | 570 (90) | 0.727 |
| ACE inhibitors/ARBs | 468 (68) | 25 (45) | 443 (70) | **< 0.001** |
| Statin | 681 (99) | 51 (93) | 630 (99) | **0.003** |
| PCSK9-i | 9 (1) | 0 (0) | 9 (1) | 1.000 |

Data are given as number (percentage), unless otherwise indicated. ACE, angiotensin-converting enzyme; ARB, angiotensin receptor blocker; BMI, body mass index; CABG, coronary artery bypass grafting; eGFR, estimated glomerular filtration rate; HDL-C, high-density lipoprotein cholesterol; IQR, interquartile range; LDL-C, low-density lipoprotein cholesterol; LVEDd, left ventricular end-diastolic diameter; LVEF, left ventricular ejection fraction; MACCE, major adverse cardiac and cerebrovascular events; MI, myocardial infarction; NSTEMI, non-ST-segment-elevation myocardial infarction; PCI, percutaneous coronary intervention; PCSK9-i, proprotein convertase subtilisin/kexin type 9 inhibitor; SBP, systolic blood pressure; SD, standard deviation; SMuRF, standard modifiable cardiovascular risk factor; and STEMI, ST-segment-elevation myocardial infarction.

During a median follow-up of 6.3 years (IQR: 4.0-8.6), 165 patients (23.9%) experienced MACCE, with an incidence rate of 4.18 events per 100 patient-years (95% CI: 3.54-4.83). Recurrent MI was the most frequent event, occurring in 70 patients (1.70 events per 100 patient-years, 95% CI: 1.30-2.11), followed by ischemia-driven revascularization in 61 patients (1.45 events per 100 patient-years, 95% CI: 1.09-1.82), all-cause death in 26 patients (0.59 events per 100 patient-years, 95% CI: 0.39-0.82), and stroke in 8 patients (0.18 events per 100 patient-years, 95% CI: 0.07-0.32).

### SMuRF Pattern

Smoking was the most prevalent SMuRF, present in 524 patients (76%), followed by hypercholesterolemia in 296 patients (43%), hypertension in 276 patients (40%), and diabetes in 83 patients (12%). Overall, 242 patients (35%) had one SMuRF, 268 patients (39%) had two SMuRFs, 102 patients (15%) had three SMuRFs, and 23 patients (3%) had all four SMuRFs. The most common combination of SMuRFs was smoking and hypercholesterolemia, observed in 113 patients (16%), followed by hypertension and hypercholesterolemia in 109 patients (16%), and the combination of smoking, hypercholesterolemia, and hypertension in 75 patients (11%) (Figure 2).

**Figure 2.**
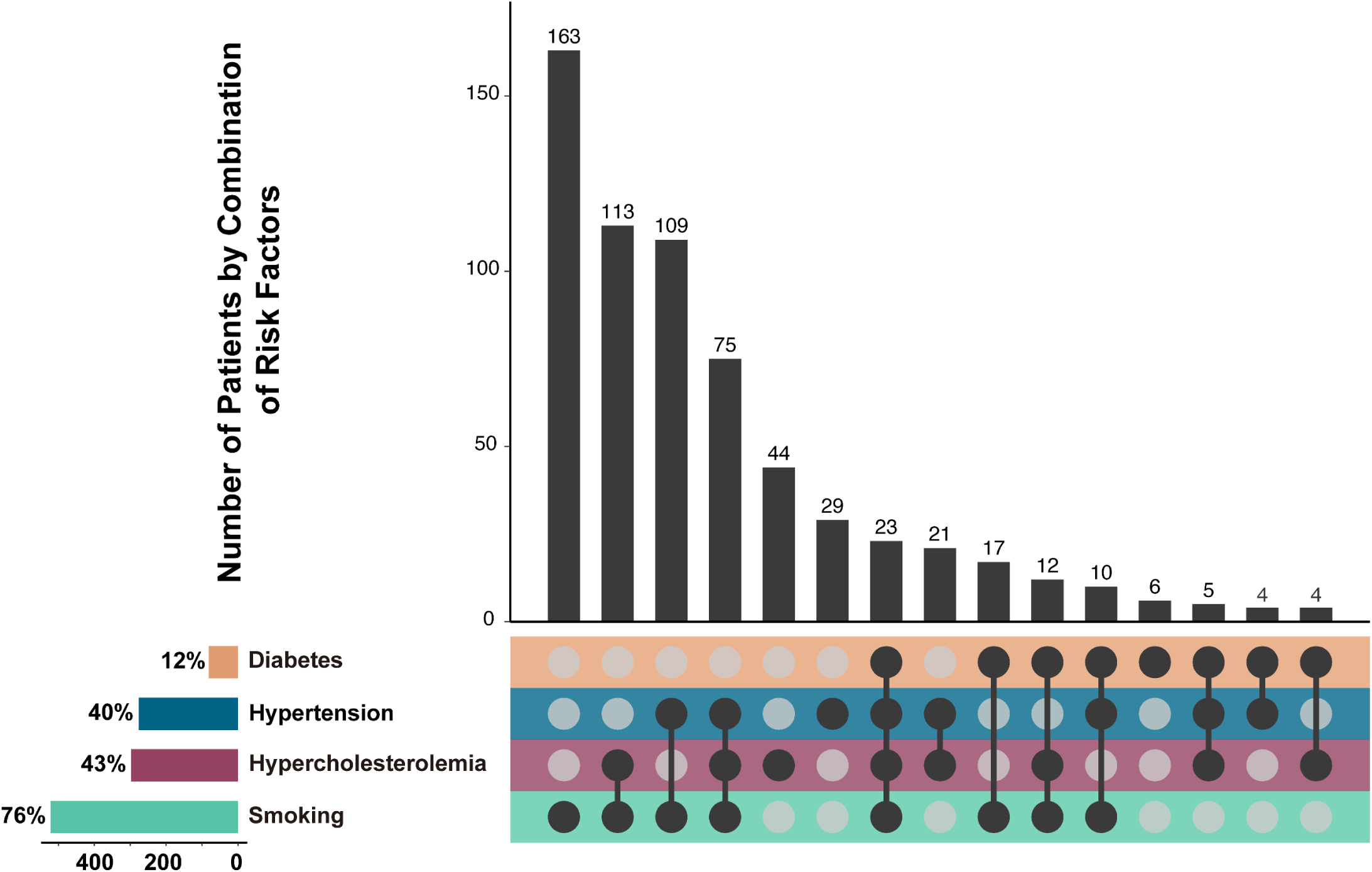
Distribution of Standard Modifiable Risk Factors. UpSet plot describing the distribution and combination of standard modifiable cardiovascular risk factors, including diabetes, hypertension, hypercholesterolemia, and smoking.

### Comparison Between SMuRF-less and SMuRF Patients

SMuRF-less patients were younger (median age: 32 years [IQR: 28.5-34] vs. 33 years [IQR: 31-34], *P*=0.014) and more likely to be female (80% vs. 96%, *P*<0.001) compared to patients with SMuRFs. Obesity was less common in the SMuRF-less group (24% vs. 46%, *P*=0.003), with a lower mean body mass index (BMI) of 25.9 ± 3.5 kg/m² compared to 28.3 ± 3.4 kg/m² in patients with SMuRFs (*P*<0.0001). The prevalence of a family history of premature CAD was similar between the groups. Established inflammatory diseases were more frequent among SMuRF-less patients (13% vs. 2%, *P*=0.003). Triglycerides level was significantly lower among SMuRF-less patients (1.5 mmol/L [IQR: 0.9, 1.9] vs. 2.0 mmol/L [IQR: 1.4, 3.0], *P*<0.001), however, high-density lipoprotein cholesterol was similar between the groups (0.9 mmol/L [IQR: 0.8, 1.1] vs. 0.9 mmol/L [IQR: 0.8, 1.0], *P*=0.150).

Coronary artery involvement was less extensive in SMuRF-less patients, with fewer left circumflex (33% vs. 46%, *P*=0.037) and right coronary artery involvement (33% vs. 54%, *P*=0.003). Fewer SMuRF-less patients underwent PCI (67% vs. 85%, *P*=0.002) and were less likely to be prescribed statins (93% vs. 99%, *P*=0.003) or ACE inhibitors/ARBs (45% vs. 70%, *P*<0.001) at discharge.

### Association of SMuRFs with MACCE

During follow-up, the incidence of new-onset diabetes was comparable across groups with different numbers of SMuRFs (*P*=0.110). Adherence to antiplatelet therapy, β-blockers, statins, and ACE inhibitors/ARBs was significantly lower among SMuRF-less patients than those with SMuRFs. Patients with 1-2 SMuRFs had lower rates of medication adherence compared to those with ≥3 SMuRFs. The rate of MACCE was significantly higher in patients with ≥3 SMuRFs (*P*=0.004). There was a trend toward higher rates of recurrent MI and ischemia-driven revascularization among patients with more SMuRFs, while the rate of all-cause death tended to be higher in the SMuRF-less group (Table 2). Similarly, cumulative hazards for MACCE were comparable between SMuRF-less patients and those with one or two risk factors but significantly higher in patients with ≥3 SMuRFs (*P*=0.003) (Figure 3).

**Figure 3.**
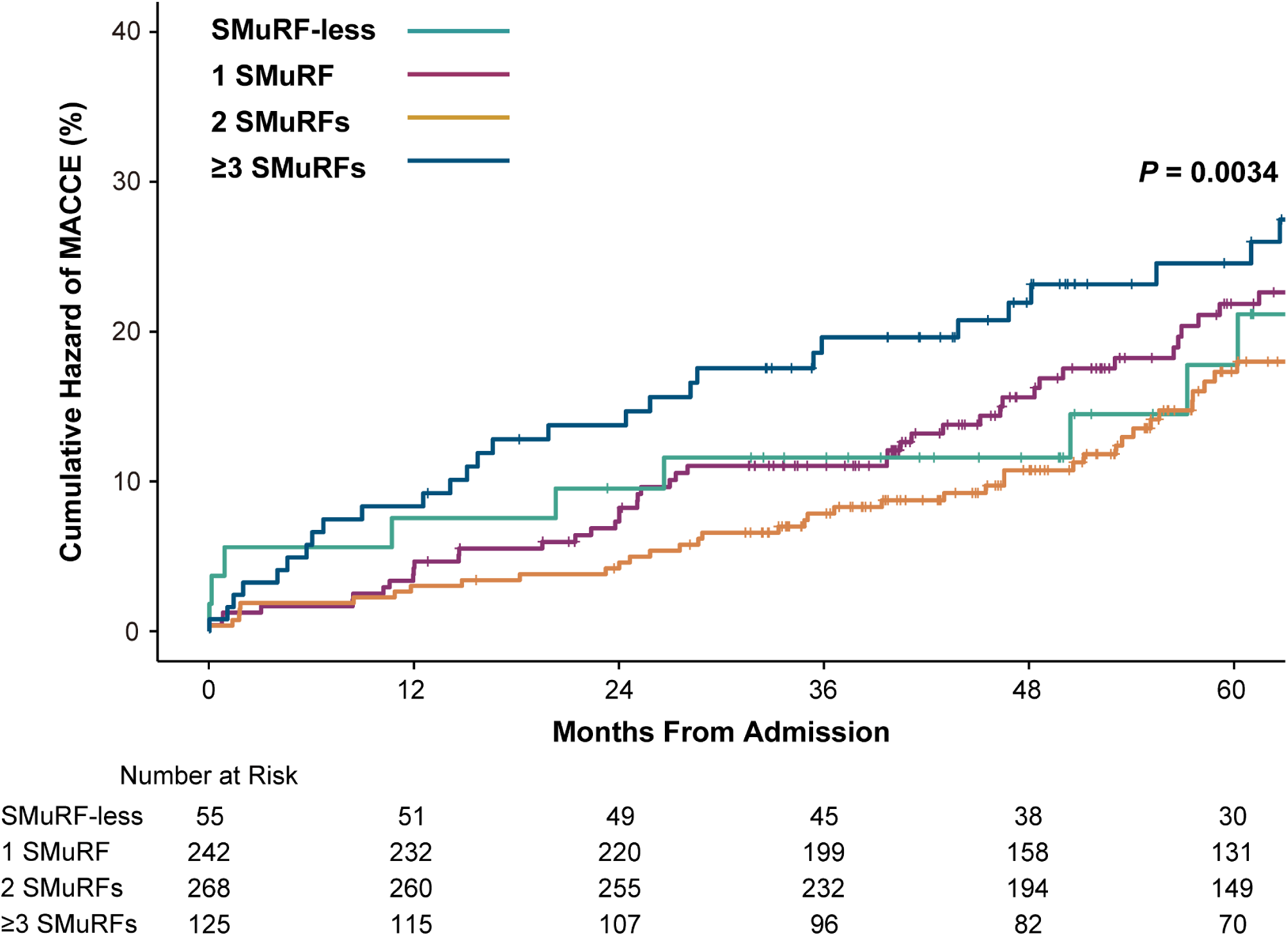
Cumulative Hazard of MACCE Stratified by Number of SmuRFs. Kaplan-Meier curves estimate cumulative event-free survival for the primary endpoint, stratified by the number of SMuRFs. MACCE, major adverse cardiac and cerebrovascular events; SMuRF, standard modifiable cardiovascular risk factor.

**Table 2.** Risk Factors, Medications, and Outcomes During Follow-up

|  | **SMuRF-less**<br>**(N = 55)** | **1 SMuRF**<br>**(N = 242)** | **2 SMuRFs**<br>**(N = 268)** | **≥3 SMuRFs**<br>**(N = 125)** | **P Value** |
| --- | --- | --- | --- | --- | --- |
| New-onset Diabetes | 5 (10) | 41 (18) | 28 (11) | 14 (11) | 0.110 |
| Current smoker | 10 (20) | 86 (37) | 123 (47) | 52 (44) | **0.002** |
| Any anti-platelet therapy | 40 (78) | 210 (91) | 238 (91) | 115 (97) | **0.001** |
| β Blockers | 32 (63) | 169 (73) | 197 (75) | 98 (83) | **0.035** |
| ACE inhibitors/ARBs | 14 (27) | 107 (46) | 123 (47) | 74 (63) | **<0.001** |
| Statin | 34 (67) | 190 (82) | 215 (82) | 107 (91) | **0.003** |
| PCSK9-i | 4 (8) | 17 (7) | 22 (8) | 13 (11) | 0.700 |
| **Outcome** |  |  |  |  |  |
| MACCE | 11 (20) | 54 (22) | 54 (20) | 46 (37) | **0.004** |
| All-cause death | 4 (7) | 10 (4) | 6 (2) | 6 (5) | 0.193 |
| Recurrent MI | 5 (9) | 22 (9) | 24 (9) | 19 (15) | 0.259 |
| Ischemia-driven<br>revascularization | 2 (4) | 18 (7) | 23 (9) | 18 (14) | 0.082 |
| Ischemic stroke | 0 (0) | 4 (2) | 1 (0.3) | 3 (2) | 0.206 |

Data are given as number (percentage), unless otherwise indicated. ACE, angiotensin-converting enzyme; ARB, angiotensin receptor blocker; MACCE, major adverse cardiac and cerebrovascular event; MI, myocardial infarction; PCSK9-i, proprotein convertase subtilisin/kexin type 9 inhibitor; and SMuRF, standard modifiable cardiovascular risk factor.

Similar to the unadjusted model (Figure S1), after adjusting for age, sex, statins, and ACE inhibitors/ARBs at discharge, hypertension (HR: 1.50; 95% CI: 1.09-2.06; *P*=0.014) and diabetes (HR: 1.67; 95% CI: 1.10-2.54; *P*=0.016) were independently associated with increased MACCE risk. The combination of hypertension and diabetes carried the highest risk (HR: 2.49; 95% CI: 1.53-4.06; *P*<0.001). In contrast, hypercholesterolemia (HR: 1.24; 95% CI: 0.91-1.68; *P*=0.177) and smoking (HR: 1.06; 95% CI: 0.72-1.55; *P*=0.776) did not independently increase MACCE risk, either alone or combined (HR: 1.20; 95% CI: 0.87-1.66; *P*=0.256). When smoking or hypercholesterolemia coexisted with either diabetes or hypertension, these combinations were strongly correlated with an increased risk of MACCE (Figure 4A). These findings remained consistent after adjustment for age, sex, family history of premature CAD, obesity, and the remaining SMuRFs (Figure 4B), except for the combination of diabetes and hypercholesterolemia, which was no longer statistically significant (HR: 1.64; 95% CI: 0.96-2.81; *P*=0.071). No interactions were detected among SMuRFs in either the adjusted or unadjusted Cox proportional hazards models (Table S1-2).

**Figure 4.**
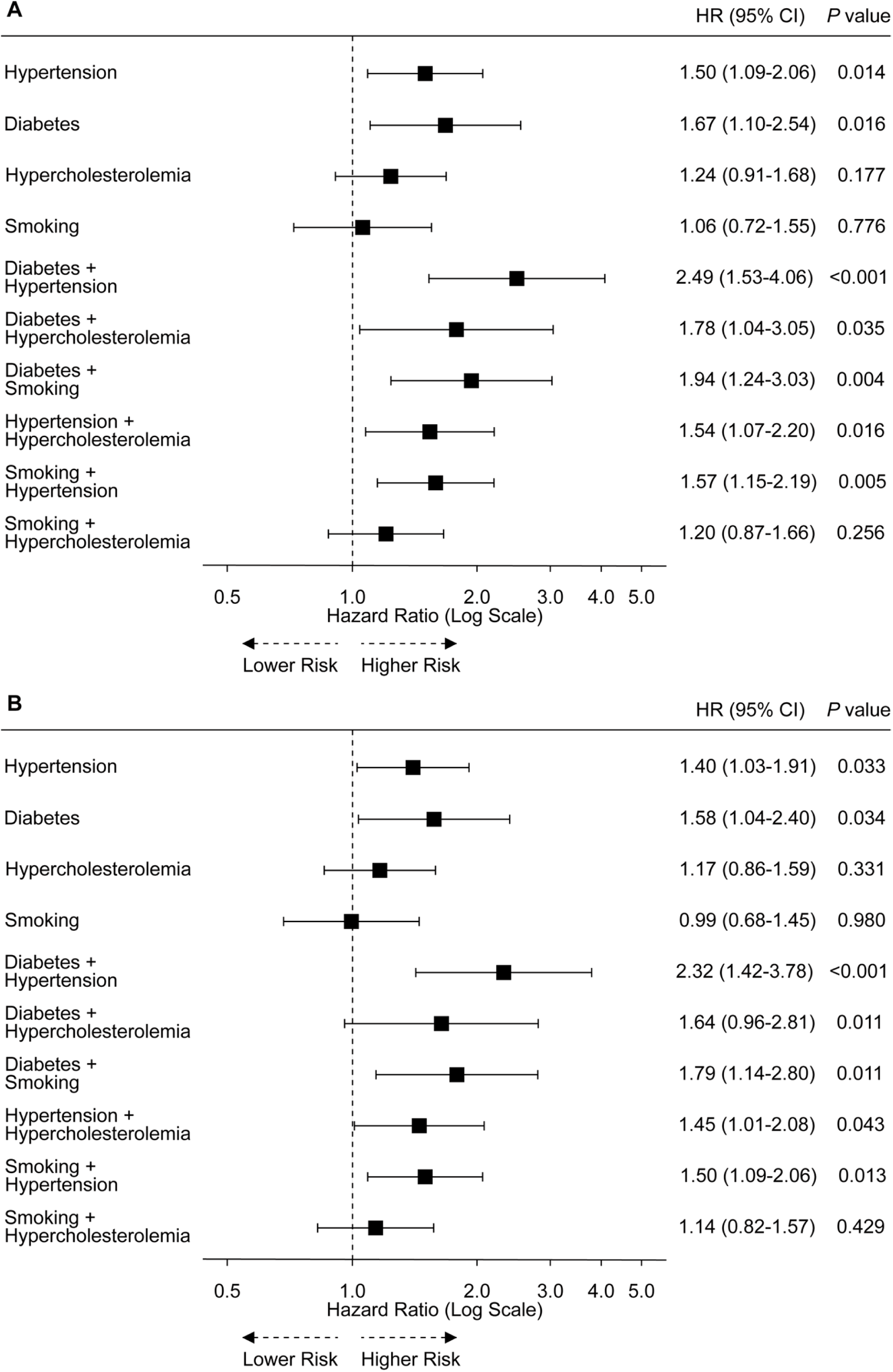
Hazard Ratio Plot from Multivariate Cox Model for MACCE of SMuRFs. Multivariate Cox Model Adjusted for (A) age, sex, β-blockers, ACE inhibitors/ARBs, and statins at discharge, and (B) age, sex, family history of premature CAD, obesity, and the remaining SMuRF. ACE indicates angiotensin-converting enzyme; ARB, angiotensin receptor blocker; MACCE, major adverse cardiac and cerebrovascular events; SMuRF, standard modifiable cardiovascular risk factor

## Discussion

Among 690 patients who experienced MI at a very young age, over 90% had at least one SMuRF, with a notably high prevalence of smoking. Increased SMuRF exposure was associated with an elevated risk of MACCE. Diabetes and hypertension, whether occurring independently or in combination, were linked to an elevated risk of MACCE. To our knowledge, this is the first study and the largest contemporary cohort to exclusively analyze SMuRF profiles in very young MI patients. Despite their younger age, our findings highlight the substantial burden of SMuRFs in this group and provide critical insights for improving secondary prevention strategies.

Traditional modifiable cardiovascular risk factors are pivotal in the pathogenesis of premature MI.^3,20^ The INTERHEART study attributed 94% of MI risk in individuals under 60 to modifiable risk factors.^21^ Similarly, the Pathobiological Determinants of Atherosclerosis in Youth (PDAY) study demonstrated that risk scores, based on factors like age, sex, lipoproteins, smoking, hypertension, obesity, and hyperglycemia, were linked to advanced atherosclerotic lesions in individuals aged 15 to 34.^22^ In line with these findings, high rates of SMuRFs were observed in patients with acute coronary syndrome (ACS), with only approximately 10% having no SMuRFs. A global meta-analysis estimated that 11.6% of ACS patients were SMuRF-less,^23^ while the Improving Care for Cardiovascular Disease in China-Acute Coronary Syndrome project (2014–2019) reported a similar rate of 11.0% in China.^24^ In a Korean STEMI cohort, Singh et al. reported a lower rate of 6.4%.^25^ However, in younger MI patients, the YOUNG-MI registry found a higher SMuRF-less rate of 17% among patients under 50 years old.^15^ Notably, the proportion of SMuRF-less patients in our cohort was lower, which we attribute to the higher prevalence of smoking compared to the 50% reported in the YOUNG-MI registry. The significant proportion of smokers in our cohort suggests that smoking may be a key driver of MI in very young populations.^26^ However, a case-control study is warranted to validate this association.

When comparing the characteristics of patients with or without SMuRF, there were two noteworthy findings. First, despite the small sample size, SMuRF-less patients were younger, more often female, and had higher rates of inflammatory diseases. The link between inflammatory disease and CAD is well-established.^27,28^ Although patients with inflammatory diseases are often comorbid with a higher prevalence of SMuRFs, such as hypertension in SLE, our younger cohort with limited SMuRF exposure supported the notion that autoantibody-mediated cardiovascular risk extends beyond SMuRFs.^29^

Second, the family history of premature CAD did not differ between SMuRF-less individuals and those with SMuRFs. Several studies have demonstrated that a family history or genetic risk score (GRS) for CAD was strongly associated with incident CAD,^30^ disease severity,^31^ and improved risk prediction.^32^ However, the overall contribution of genetic predisposition to CAD incidence remains modest or neutral. For instance, a Finnish study using a GRS of 28 CAD risk variants found only a 12% reclassification when added to conventional risk factors.^32^ Similarly, a Danish cohort using a GRS of 45 variants found that genetic risk did not significantly enhance risk stratification beyond the traditional European SCORE. Also, the association between GRS and incident MI was insignificant in individuals under 45 years old.^33^ Moreover, the risk of CAD in individuals with a high genetic predisposition could be reduced by up to 50% through positive lifestyle changes.^34^ Thus, addressing traditional risk factors and promoting lifestyle interventions remain essential for managing the risk of premature CAD.^35^

Although data on SMuRF-less patients is limited, the current consensus recommends applying secondary prevention strategies to all patients, including those without SMuRFs.^18,36^ We observed a lower rate of prescribing ACE inhibitors/ARBs and statins at baseline and during follow-up in SMuRF-less patients. Additionally, patients with one SMuRF had the highest rate of new-onset diabetes during follow-up. The poorer adherence to secondary prevention may partly explain why the number of SMuRFs showed only a partial dose-response relationship with MACCE in our cohort, with event rates comparable between SMuRF-less patients and those with one or two risk factors.

Diabetes and hypertension were the two most significant outcome-related factors in our study. The Partners YOUNG-MI registry reported that diabetes was associated with a 2.68-fold increased risk of cardiovascular mortality.^37^ The DDCD study also identified baseline diabetes as the only independent predictor of a first ischemic recurrence.^13^ In the Appraisal of risk Factors in young Ischemic patients Justifying aggressive Intervention (AFIJI) study, baseline diabetes was linked to a 1.75-fold increased risk of a first recurrent event.^12^ Hypertension demonstrated a weaker association with recurrences in previous premature MI studies and was associated only with multiple recurrences in the AFIJI study.^12^ However, in studies across all age groups, hypertension is independently associated with cardiac death, recurrent MI, heart failure, stroke, and reduced improvement in LV systolic function after MI.^38,39^ Whilst the underlying mechanisms remain unclear, an observed mechanism might involve a propensity to myocardial haemorrhage, which is mediated by hypertension-induced microvascular injury.^39^

Smoking and hypercholesterolemia were not independently associated with prognosis in our cohort. In the AFIJI study, however, persistent smoking emerged as the strongest predictor of a first recurrent event, with smoking rates at 35% among patients with multiple recurrences, compared to just 1.5% in those without ischemic recurrences.^12^ Consistently, the Partners YOUNG-MI registry reported that nearly 25% of premature MI patients continued smoking, which was associated with a 50% increased risk of death compared to those who quit within a year.^26^ A possible explanation for the observed disparity in our cohort is that our patients were 5–10 years younger, with shorter exposure to risk factors, leading to a reduced impact on long-term outcomes. While baseline hypercholesterolemia did not affect long-term outcomes in our study, a higher initial statin dose post-MI is linked to better outcomes,^40^ underscoring the importance of optimal LDL-C control.

## Limitations

This study provided insights into the characteristics and outcomes of very young MI patients within a contemporary Chinese cohort. However, several limitations warrant consideration. First, the retrospective design limits our ability to establish definitive causal relationships between risk factors and outcomes. Second, the study was conducted within two tertiary medical centers in Beijing, China. This geographic limitation may affect the generalizability of the findings to broader populations. Future studies involving diverse geographic regions would be valuable for validating these findings. Third, recall bias and missed intervals may have occurred due to our follow-up strategy, which included both clinical visit records at our centers and phone calls to track events not captured at our centers. Fourth, while this study focused on standard modifiable risk factors (SMuRFs), other risk factors may influence young MI patients. Future research should explore how non-SMuRF factors contribute to cardiovascular outcomes, particularly in younger demographics.

## Conclusion

Young MI patients have a high burden of standard modifiable risk factors, with diabetes and hypertension significantly increasing long-term risk.

## Data Availability

The data supporting the findings of this study are available upon reasonable request. Interested researchers can contact the corresponding author via email at.

## Acknowledgments

T-SQ and Z-XL contributed equally to data collection, analysis, and the writing of the paper. L-YB, H-TY, C-YX, and L-JY contributed to data collection. T-SQ, Z-XL, and R-YP contributed to patient follow-up. F-ZJ and R-YP contributed to the patient’s clinical care, therapy plan, and critical review of the paper.

## Supplement materials

**Figure S1.** Unadjusted Hazard Ratio Plot from Multivariate Cox Model for MACCE of SMuRFs

**Table S1.** Unadjusted Cox Proportional Hazards Model for SMuRF Interaction

**Table S2.** Adjusted Cox Proportional Hazards Model for SMuRF Interaction

## Non-standard Abbreviations and Acronyms Abbreviation/Acronym Expanded Form

ACE inhibitor: Angiotensin-Converting Enzyme Inhibitor
AFIJI: Appraisal of Risk Factors in Young Ischemic Patients Justifying Aggressive Intervention
ARB: Angiotensin Receptor Blocker
DDCD: Duke Databank for Cardiovascular Disease
GRS: Genetic Risk Score
MACCE: Major Adverse Cardiovascular and Cerebrovascular Events
PCSK9-i: Proprotein convertase subtilisin/kexin type 9 inhibitor
PDAY: Pathobiological Determinants of Atherosclerosis in Youth
SMuRF: Standard Modifiable Risk Factor

